# Central corneal thickness and the risk of primary open-angle glaucoma: a Mendelian randomization mediation analysis

**DOI:** 10.1101/2023.11.28.23299139

**Authors:** Andreas Katsimpris, Sebastian-Edgar Baumeister, Hansjörg Baurecht, Andrew J Tatham, Michael Nolde

**Author notes:** shared last author. Corresponding authors: Andreas Katsimpris, Department of Ophthalmology, Aberdeen Royal Infirmary, Scotland, AB25 2ZN, UK, ORCID ID: https://orcid.org/0000-0002-5805-105X, Michael Nolde, Institute of Health Services Research in Dentistry, University of Münster, Germany, ORCID ID: https://orcid.org/0000-0001-6893-7367.

## Abstract

The association of central corneal thickness (CCT) with primary open-angle glaucoma (POAG) remains uncertain. Although, several observational studies assessing this relationship, have reported an inverse association between CCT and POAG, this could be the result of collider bias. In this study, we leveraged human genetic data to assess through Mendelian randomization (MR) the effect of CCT on POAG risk, and whether this effect is mediated by intraocular pressure (IOP) changes. We used 24 single-nucleotide polymorphisms (SNPs) associated with CCT (P-value < 5×10^-8^) from a genome-wide association study (GWAS) (N = 17,803) provided by the International Glaucoma Genetics Consortium and 53 SNPs associated with IOP (P-value < 5×10^-8^) from a GWAS of UK Biobank (UKBB) (N = 97,653). We related these instruments with POAG using a GWAS meta-analysis of 8,283 POAG cases and 753,827 controls from UKBB and FinnGen. MR analysis suggested a positive association between CCT and POAG (odds ratio of POAG per 50μm increase in CCT: 1.38; 95% confidence interval: 1.18 to 1.61; p-value < 0.01). MR mediation analysis showed that 28.4% of the total effect of CCT on POAG risk was mediated through changes in IOP. The primary results were consistent with estimates of pleiotropy-robust MR methods. In conclusion, contrary to most observational studies, our results support a positive effect of CCT on the risk of POAG.

## Introduction

Primary open-angle glaucoma (POAG) ranks as the primary cause of irreversible blindness worldwide, with projections indicating a significant and growing impact in the future [1]. Identification of recognized risk factors for POAG, such as elevated intraocular pressure (IOP), family history of POAG and non-White ethnicity in individuals can lead to early detection of POAG through screening initiatives and mitigate vision impairment [2].

The role of central corneal thickness (CCT) as a potential risk factor for POAG remains uncertain, with ongoing debate regarding its clinical significance in the diagnosis and management of the condition [3]. Lower CCT has been postulated to be spuriously associated with higher risk of POAG, since CCT can artificially influence IOP measurements [4]. Moreover, the inverse association between CCT and POAG has been reported mainly by observational studies, which either adjusted for IOP in their analyses, thereby treating IOP like a confounder [5,6], or selected their participants based on measured IOP [7]. This can result in a spurious inverse association of CCT with POAG, as a result of collider bias [8]. Collider bias in a study can occur after controlling for a variable that is a common effect of the exposure and the outcome. The variable that is caused by both the exposure and the outcome is termed a “collider”, and controlling for this variable either by study design or statistical analysis can create spurious associations between the exposure and the outcome of interest. In studies assessing the association between CCT and POAG, collider bias can occur when measured IOP is controlled, either in study design or statistical analysis, since measured IOP is causally affected by both true IOP and CCT (Figure 1).

**Figure 1.**
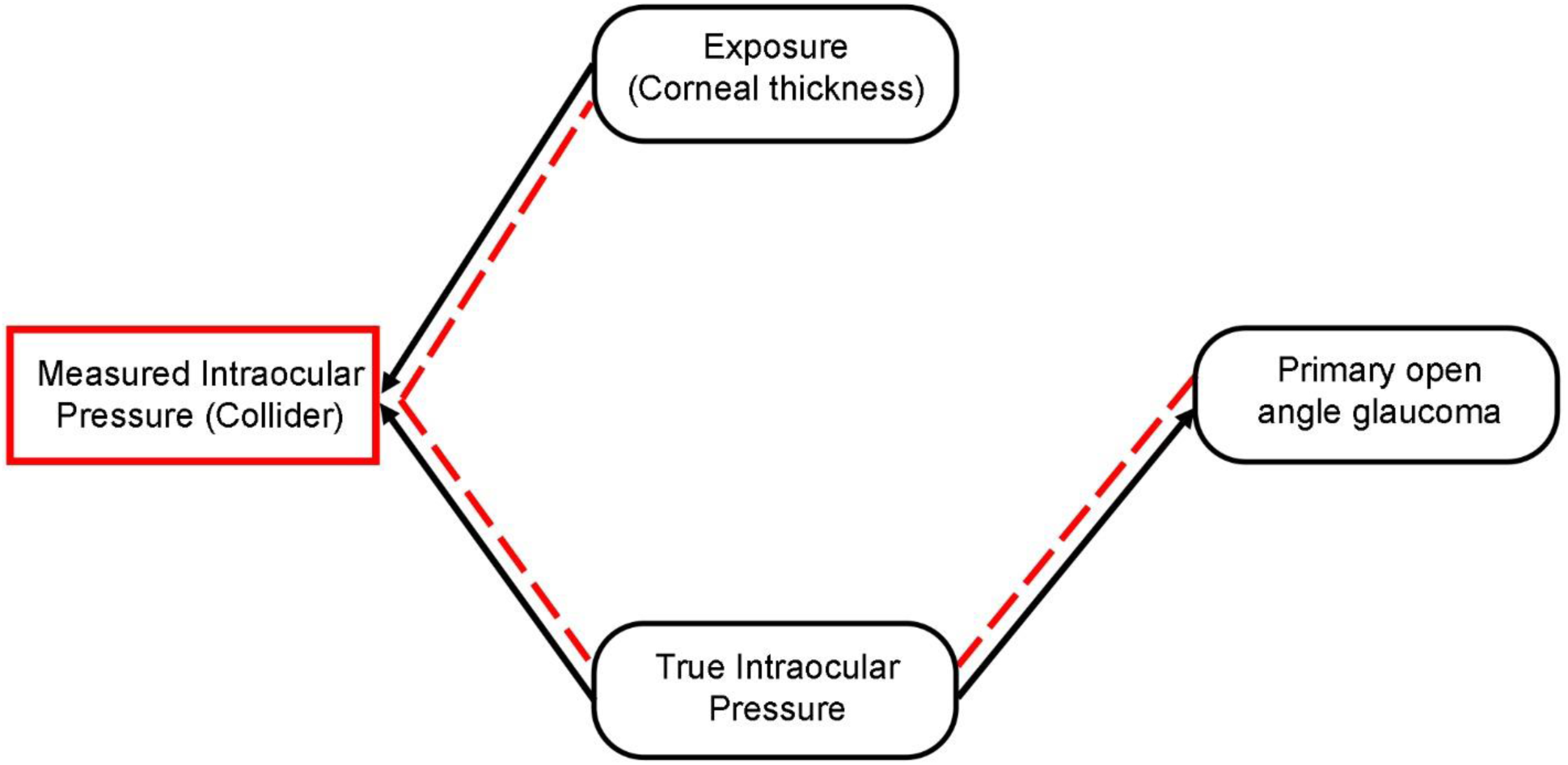
Directed acyclic graph showing the existence of collider bias when assessing the association of central corneal thickness (CCT) with primary open angle glaucoma (POAG). Measured IOP is a collider, since both CCT and true IOP are causally associated with it (black arrows), In cases of adjustment for measured IOP in the analysis or stratification of the analysis based on measured IOP or selection of the study participants based on measured IOP, a spurious relationship (dashed red line) will occur between CCT and POAG via true IOP, even if no true causal association between CCT and POAG exists (there is no arrow connecting CCT with POAG).

One method to assess the existence of a causal relationship between CCT and POAG, is Mendelian randomization (MR), a type of instrumental variable analysis, where genetic variants from genome-wide association studies (GWAS) are used as instruments [9]. It is noteworthy that a recent MR study found a marginally nonsignificant positive association between CCT and POAG [10]. This suggests that the causal relationship between CCT and POAG may follow an opposite direction compared to what has been observed in most observational studies.

Considering that the lack of statistical significance of the association estimate from the recent MR study may have been due to the relatively small sample size of the GWAS used, we employed MR in our current study, utilizing larger GWAS datasets of POAG to assess the existence of a causal association between CCT and POAG. Moreover, we conducted a two-step MR for mediation analysis [11], to further investigate the proportion of the effect of CCT on POAG mediated through IOP changes.

## Materials and Methods

### Study design

MR employs genetic variants, typically single-nucleotide polymorphisms (SNPs), as instrumental variables to assess the effect of modifiable risk factors on disease risk [9]. These genetic variants are randomly allocated at conception, akin to a natural randomized controlled trial, reducing susceptibility to confounding and reverse causation biases [11]. In our study, we conducted a two-sample MR using summary statistics from GWAS for CCT [12] and POAG [13,14], in order to assess the effect of CCT on POAG risk. Additionally, we performed mediation analysis with two-step MR [11] to further explore the proportion of the effect of CCT on POAG mediated through IOP changes. We followed the STROBE-MR guidelines [15] and “Guidelines for performing Mendelian randomization investigations” [16] and we have not pre-registered the study protocol.

### Data sources

We retrieved summary data from the largest GWAS to date for CCT [10] comprising 17,803 individuals of European descent, from the International Glaucoma Genetics consortium IGGC. CCT in individual cohorts was assessed with ultrasound pachymetry or corneal topography and was measured in μm. Genotyping and imputation methods of the GWAS have been described elsewhere [10]. Summary statistics for POAG were retrieved from two GWAS: 1) the FinnGen consortium database (R8 release), which included 6,785 POAG cases and 349,292 controls [13], 2) the UK Biobank (UKBB) cohort, which included 1,498 POAG cases and 404,535 controls [14]. In both GWAS participants were of European descent and POAG cases met the criteria for a diagnosis of POAG based on the International Classification of Diseases, Ninth Revision (ICD-9) or International Statistical Classification of Diseases, Tenth Revision (ICD-10) code. Summary statistics for corneal-compensated IOP (IOPcc) were also retrieved from the UKBB cohort [14]. More specifically, a sub-sample of 97,653 participants from the UKBB underwent ophthalmic assessment including IOPcc assessment in millimeters of mercury (mmHg) using an Ocular Response Analyzer non-contact tonometer. Our chosen IOP phenotype was IOPcc since it was designed to account for corneal biomechanical properties and has also been utilized in prior GWAS for IOP [17]. Genotyping, quality control and imputation methods of the GWAS have been described elsewhere [13,18].

### Selection of genetic variants as instrumental variables

We chose SNPs from the CCT GWAS that reached genome-wide significance (P-value < 5*10^-8^) after clumping for linkage disequilibrium (LD) at r^2^ < 0.001 over a 10mb window. Utilizing the MR-Steiger directionality test, we determined the causality direction between CCT and POAG [19]. SNPs more strongly correlated with the outcome than the exposure were excluded, along with those showing significant influence in the funnel plots and scatter plots. Ultimately, 24 SNPs associated with CCT were selected as instrumental variables. Moreover, by summing the coefficients of determination (R^2^) obtained from the associations between the selected SNPs and CCT, we calculated the percentage of variability in CCT that can be accounted for by the selected 24 SNPs. In a similar way, we selected 53 genetic variants from the IOP GWAS for our MR mediation analysis.

### Statistical analysis

The SNP-POAG association estimates of the selected SNPs were extracted from a meta-analysis of the FinnGen and UKBB GWAS, that we performed using the inverse-variance weighted (IVW) fixed effect approach. Following data harmonization, which involved filtering SNPs based on HapMap3 [20], excluding strand-ambiguous ones, and aligning effect sizes, we computed Wald ratios. These ratios were obtained by dividing the per-allele logarithm of odds ratio (logOR) for each SNP from the meta-analyzed POAG GWAS by its corresponding logOR from the GWAS for CCT. The cumulative effect of CCT on POAG risk was then estimated through a multiplicative random effects IVW meta-analysis of the Wald ratios [21].

We conducted a univariable two-sample MR using summary-level statistics from GWAS available for CCT and POAG. The two-sample MR approach relies on three fundamental assumptions: (1) the genetic instruments should be reliably associated with the risk factor under investigation (“relevance” assumption), (2) the genetic instruments should not be associated to factors that might confound the association between the exposure and outcome (“exchangeability” assumption), and (3) the genetic instruments are not associated with the outcome other than via the risk factor of interest (“exclusion restriction” assumption) [22,23]. To fulfill the “relevance” assumption, we ensured that the selected SNPs as instrumental variables reached genome-wide significance (P-value < 5*10^-8^). Additionally, we assessed instrument strength by calculating the F-statistic of the selected genetic instruments as well as the proportion of exposure variance they explain. [24]. While the “exchangeability” and “exclusion restriction” assumptions cannot be definitively proven, we conducted sensitivity analyses to detect potential violation of the assumptions underlying MR. Possible violations may arise from horizontal pleiotropy, where genetic variants impact outcomes through pathways unrelated to the investigated exposure. Thus, we employed PhenoScanner [25] to assess associations between our selected genetic instruments and traits that could potentially confound our analysis. If pleiotropic pathways were detected, we utilized multivariable MR to account for these effects [26]. Furthermore, we examined each selected SNP and its proxies for associations with known POAG risk factors, assessed heterogeneity among the chosen genetic variants through the Cochran Q heterogeneity test and I_GX_^2^ [23] to detect pleiotropy, and conducted MR Egger regression [23] and pleiotropy-robust methods [27] (penalized weighted median, IVW radial regression, and MR-Pleiotropy Residual Sum and Outlier (MR-PRESSO)) to assess directional pleiotropy. To determine if the IVW estimate was influenced by a single SNP, we conducted a leave-one-out analysis.

For assessing the effect of CCT on POAG that is mediated through IOP (indirect effect), we conducted a two-step MR for mediation analysis [11]. In this method 2 MR estimates are calculated 1) the causal effect of CCT on the IOP using a univariable MR model and 2) the causal effect of the IOP on POAG using a multivariable MR model adjusted for CCT. These 2 estimates are then multiplied together to estimate the indirect effect of CCT on POAG that is mediated through IOP. The total effect of CCT on POAG was also calculated and in all these mediation MR analyses we used only the POAG GWAS from the FinnGen cohort, in order to avoid overlap with the UKBB GWAS for IOP. Additionally, we calculated the proportion of the total effect of CCT on POAG explained by the mediator (IOP), by dividing the indirect effect of CCT on POAG by the total effect. The delta method was used to estimate 95% confidence intervals (95%CI) for the indirect effect and the proportion mediated [28]. MR for mediation requires SNPs that have been selected as instruments for the exposure and mediator to be independent [11], so we ensured our selected SNPs from the CCT and IOP GWAS to be non-overlapping.

All MR estimates for the associations of CCT with IOP and POAG were multiplied by 50, representing the change in log odds of POAG or units of IOP per 50μm increase in CCT. All analyses were performed with R version 4.2.1 [29] using the MendelianRandomization, TwoSampleMR, MVMR and MR-PRESSO packages.

## Results

### Effect of central corneal thickness on primary open-angle glaucoma

The selected 24 SNPs from the CCT GWAS explained 7.55% of the variance in CCT and the F-statistics for all SNPs were ≥30.87 (Supplementary Table S2). We found a positive effect of the genetically predicted CCT on POAG risk using the IVW method (OR=1.38 per 50μm increase in CCT; 95%CI = 1.18 to 1.61; P-value < 0.01) (Figure 2 and Supplementary Figure S2). The estimates from the pleiotropy-robust methods were consistent with the estimate from the IVW analysis (Figure 2). None of our instrumental SNPs were associated with POAG risk factors (Supplementary Table S3) and, thus, we did not perform multivariable MR to adjust for correlated horizontal pleiotropy.

**Figure 2.**
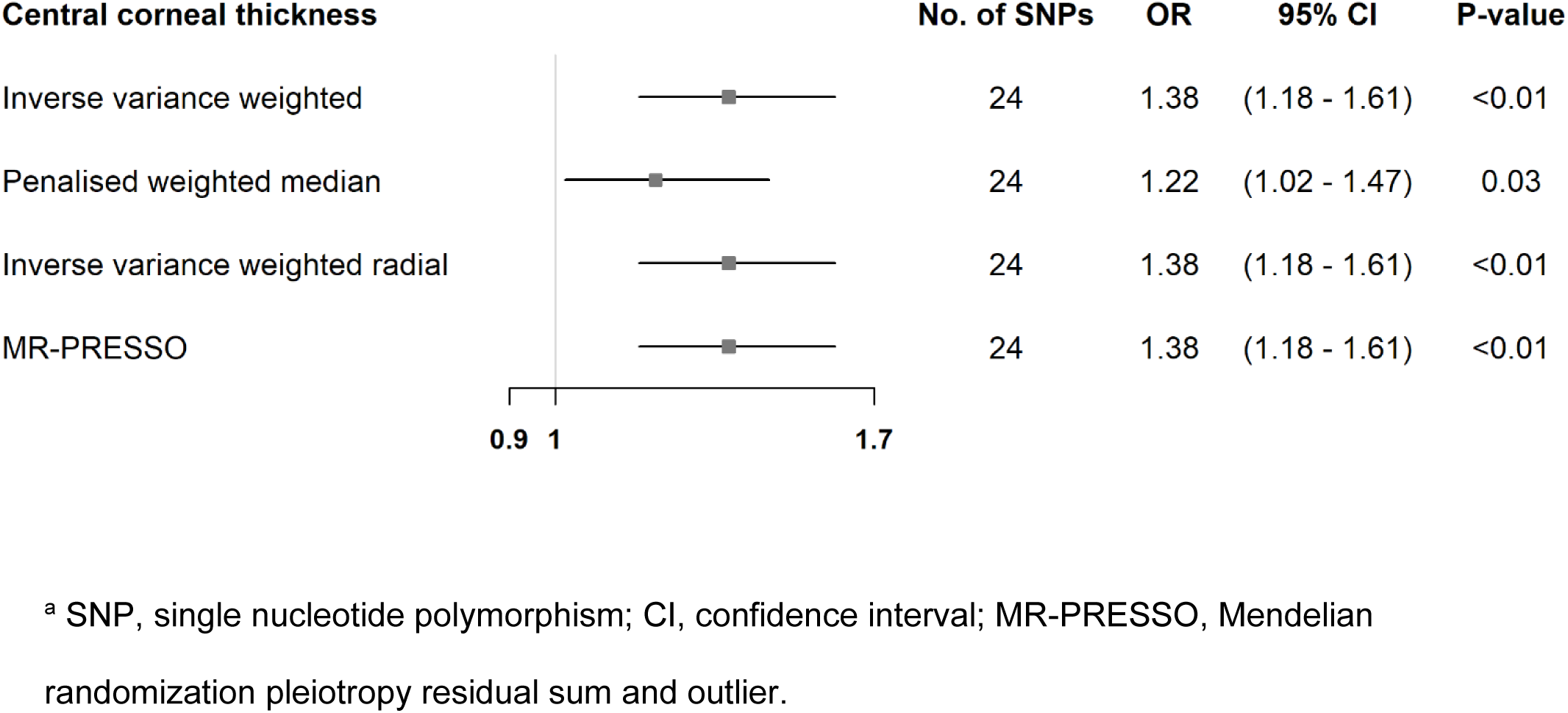
Mendelian randomization estimates for the effect of central corneal thickness on primary open-angle glaucoma. Estimates are reported as changes in odds of primary open-angle glaucoma per 50 μm increase in central corneal thickness ^a^.

We found evidence of heterogeneity among Wald ratios for CCT with POAG (Supplementary Table S4), with a Cochran’s Q heterogeneity test value of 40.62 (p-value = 0.013). However, the intercepts from the MR-Egger analyses did not deviate from zero, thus, no directional pleiotropy was present (Supplementary Table S4). The leave-one-SNP-out analyses identified no SNPs with high influence on the IVW estimates for our exposures (Supplementary Table S5).

### Mediation analysis

An illustration of the MR mediation analysis can be seen in Figure 3. Because here we used only one GWAS for POAG the estimate of the total effect of CCT on POAG was slightly different (OR: 1.50 per 50μm increase in CCT; 95%CI = 1.27 to 1.77; P-value < 0.01, Figure 2B and Supplementary Table S6). 28.4% (95%CI: 0 to 60%) of the total effect of CCT on POAG was mediated through changes in IOP.

**Figure 3.**
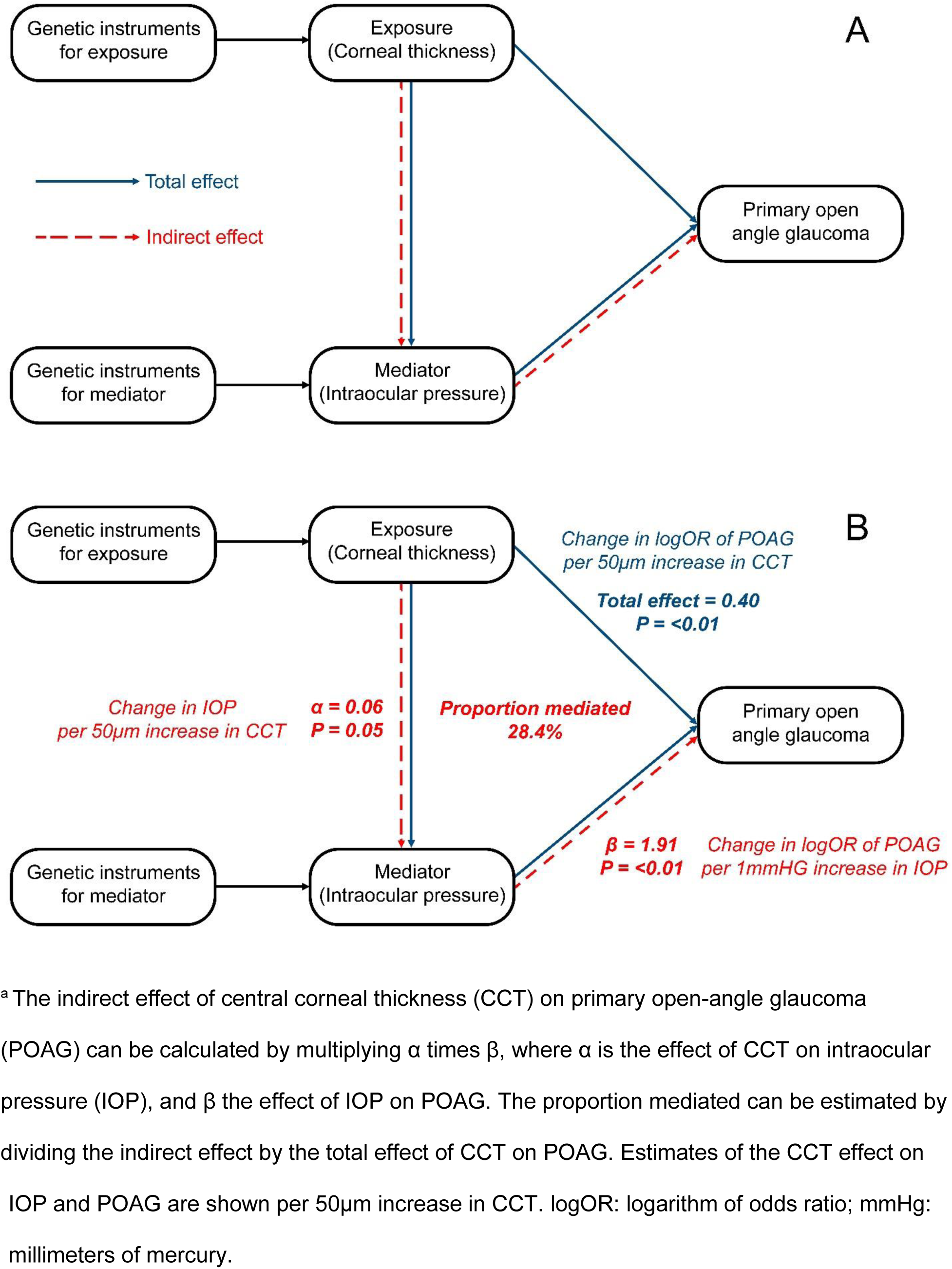
Directed acyclic graphs of the mediation analysis with Mendelian randomization ^a^.

## Discussion

In this two-sample MR, we utilized genetic data to assess the association between CCT and the risk of POAG. Moreover, we conducted MR mediation analysis to assess the proportion of the CCT effect on POAG that is mediated through IOP changes. Contrary to most of the observational studies in the literature, we found evidence of a positive causal association between CCT and POAG.

The first landmark glaucoma study to suggest that thinner corneas are associated with the development of POAG was the Ocular Hypertension Treatment Study [7]. (OHTS). In this prospective study they found an unadjusted and adjusted hazard ratio (HR) for POAG of 1.88 (95%CI = 1.55 to 2.29) and 1.71 (95%CI = 1.40 to 2.09), respectively, per 40μm decrease in CCT. However, all the OHTS study participants were individuals with ocular hypertension (IOP > 21mmHg), and since higher CCT can result in higher IOP measurement readings [30], this led to a selection of a cohort with high CCT values (93% of the total participants had CCT higher than 526 mm). As a result, individuals in the OHTS with high CCT might actually have lower true IOP which may have caused this false inverse association between CCT and POAG conversion in the OHTS due to selection bias.

Additionally, two other landmark glaucoma studies, the Los Angles Latino Eye study (LALES) [5] and Early Manifest Glaucoma Trial (EMGT) [6], have found an inverse association between CCT and POAG, but only in their confounder-adjusted analyses. In the univariate analysis of baseline factors predicting the development of POAG in the LALES the OR for POAG per 40μm decrease in CCT was 1.16 (95%CI: 0.90 to 1.50, p-value = 0.25), while in the multivariable analysis, that adjusted for IOP, this association estimate became marginally statistically significant with an OR for POAG of 1.30 (95%CI: 1.00 to 1.70, p-value = 0.05). Similarly, in the EMGT, the univariable analysis showed no association between CCT and POAG per 40μm decrease in CCT (HR: 1.23, 95%CI: 0.90–1.68, p-value = 0.188). However, in an analysis stratified on baseline IOP, a statistically significant 42% increase in POAG risk was found per 40μm decrease in CCT (HR: 1.42, 95%CI: 1.00 to 1.92, p-value = 0.02), only in patients with higher baseline IOP.

The fact that in these three landmark glaucoma studies a significant inverse association occurred only after either adjusting for IOP in their analyses or stratifying their analysis based on IOP or selecting their participants based on IOP, suggests the presence of collider bias [31]. (Figure 1). Selection bias can be considered a form of collider bias, where controlling of the collider happens during sampling of the study participants [32]. This could be the case for the presence of an inverse association between CCT and POAG in the OHTS, since participants were selected based on measured IOP (collider), and both true IOP (outcome) and CCT (exposure) are causally associated with it. In LALES [5] and EMGT [6], the significant association between CCT and POAG seems to occur due to collider bias after adjusting for measured IOP in their statistical analyses. The presence of collider bias on the CCT-POAG association has also extensively been investigated by Khawaja et al. [8], where in their simulated studies CCT was significantly associated with POAG only when adjusted for measured IOP or when participants were selected on measured IOP.

In contrast to the observational studies mentioned above, a recent two-sample MR study [10] found that genetic predisposition to higher CCT is associated with higher risk of POAG, similarly to our results, but this association was marginally not significant (OR for POAG: 1.20 per 50μm increase in CCT, 95%CI: 0.97 to 1.47, p-value = 0.09). The POAG GWAS that they used included 63,412 participants (4,986 POAG cases and 58,426 controls). We were able to detect a statistically significant association between CCT and POAG, because the combined GWAS data for POAG were much larger and consisted of 762,210 participants (8,283 POAG cases and 753,827 controls). Despite the significant heterogeneity among the Wald ratios for our selected SNPs, no directional pleiotropy, that could lead to biased estimates, was evident [23].

The key strength of this study was the large sample size of the combined GWAS for POAG, which increased the power of our study. Moreover, the association estimates from the pleiotropy-robust methods were consistent with the IVW estimate and did not indicate any model violations. Additionally, our mediation analysis with MR allowed for assessment of the causal pathway between CCT and POAG. However, some limitations need to be taken into account. First, our outcome of interest was POAG so the effect of CCT on other types of glaucoma (e.g., primary angle-closure glaucoma) were not assessed. Second, our MR models assumed a linear relationship between CCT and POAG and no interaction between these two factors.

In conclusion, contrary to most observational studies, our data provided evidence for a positive association of CCT with POAG, with almost one third of the CCT effect being mediated through IOP changes. Triangulation of evidence from different types of research studies, with different key sources of bias, is warranted to confirm these results.

## Author contributions

KA, BSE and NM contributed to the study conception and design, drafted the manuscript and analyzed the data. All authors critically revised the manuscript for important intellectual content, provided administrative, technical, or material support and approved the final version.

## Data availability

The summary statistics for the central corneal thickness GWAS are available at https://datashare.ed.ac.uk/handle/10283/2976 (access date: 2023/08/17). The primary open-angle glaucoma summary statistics for the FinnGen GWAS are available at https://www.finngen.fi/en/access_results (access date: 2023/08/17). The primary open-angle glaucoma and intraocular pressure summary statistics for the UKBB GWAS are available at https://pan.ukbb.broadinstitute.org (access date: 2023/08/17)

## Additional information

Competing Interests: None declared

## Acknowledgments

We want to acknowledge the participants and investigators of the FinnGen and UK Biobank studies.

## References

1 Tham, Y. C. et al. Global prevalence of glaucoma and projections of glaucoma burden through 2040: a systematic review and meta-analysis. Ophthalmology 121, 2081–2090, doi:10.1016/j.ophtha.2014.05.013 (2014).

2 Weinreb, R. N. et al. Primary open-angle glaucoma. Nat Rev Dis Primers 2, 16067, doi:10.1038/nrdp.2016.67 (2016).

3 Sng, C. C., Ang, M. & Barton, K. Central corneal thickness in glaucoma. Curr. Opin. Ophthalmol. 28, 120–126, doi:10.1097/icu.0000000000000335 (2017).

4 Medeiros, F. A. & Weinreb, R. N. Is corneal thickness an independent risk factor for glaucoma? Ophthalmology 119, 435–436, doi:10.1016/j.ophtha.2012.01.018 (2012).

5 Jiang, X. et al. Baseline risk factors that predict the development of open-angle glaucoma in a population: the Los Angeles Latino Eye Study. Ophthalmology 119, 2245–2253, doi:10.1016/j.ophtha.2012.05.030 (2012).

6 Leske, M. C. et al. Predictors of long-term progression in the early manifest glaucoma trial. Ophthalmology 114, 1965–1972, doi:10.1016/j.ophtha.2007.03.016 (2007).

7 Gordon, M. O. et al. The Ocular Hypertension Treatment Study: Baseline Factors That Predict the Onset of Primary Open-Angle Glaucoma. Arch. Ophthalmol. 120, 714–720, doi:10.1001/archopht.120.6.714 (2002).

8 Khawaja, A. P. & Jansonius, N. M. Potential for Collider Bias in Studies Examining the Association of Central Corneal Thickness With Glaucoma. Invest. Ophthalmol. Vis. Sci. 63, 3–3, doi:10.1167/iovs.63.12.3 (2022).

9 Sanderson, E. et al. Mendelian randomization. Nature Reviews Methods Primers 2, 6, doi:10.1038/s43586-021-00092-5 (2022).

10 Choquet, H. et al. A multiethnic genome-wide analysis of 44,039 individuals identifies 41 new loci associated with central corneal thickness. Commun Biol 3, 301, doi:10.1038/s42003-020-1037-7 (2020).

11 Carter, A. R. et al. Mendelian randomisation for mediation analysis: current methods and challenges for implementation. Eur. J. Epidemiol. 36, 465–478, doi:10.1007/s10654-021-00757-1 (2021).

12 Iglesias, A. I. et al. Cross-ancestry genome-wide association analysis of corneal thickness strengthens link between complex and Mendelian eye diseases. Nat Commun 9, 1864, doi:10.1038/s41467-018-03646-6 (2018).

13 Kurki, M. I. et al. FinnGen provides genetic insights from a well-phenotyped isolated population. Nature 613, 508–518, doi:10.1038/s41586-022-05473-8 (2023).

14 Pan-UKB team. https://pan.ukbb.broadinstitute.org. 2020.

15 Skrivankova, V. W. et al. Strengthening the reporting of observational studies in epidemiology using mendelian randomisation (STROBE-MR): explanation and elaboration. BMJ 375, n2233, doi:10.1136/bmj.n2233 (2021).

16 Burgess, S. et al. Guidelines for performing Mendelian randomization investigations: update for summer 2023 [version 3; peer review: 2 approved]. Wellcome Open Research 4, doi:10.12688/wellcomeopenres.15555.3 (2023).

17 Simcoe, M. J., Khawaja, A. P., Hysi, P. G. & Hammond, C. J. Genome-wide association study of corneal biomechanical properties identifies over 200 loci providing insight into the genetic etiology of ocular diseases. Hum. Mol. Genet. 29, 3154–3164, doi:10.1093/hmg/ddaa155 (2020).

18 Bycroft, C. et al. The UK Biobank resource with deep phenotyping and genomic data. Nature 562, 203–209, doi:10.1038/s41586-018-0579-z (2018).

19 Hemani, G., Tilling, K. & Davey Smith, G. Orienting the causal relationship between imprecisely measured traits using GWAS summary data. PLoS Genet. 13, e1007081, doi:10.1371/journal.pgen.1007081 (2017).

20 Frazer, K. A. et al. A second generation human haplotype map of over 3.1 million SNPs. Nature 449, 851–861, doi:10.1038/nature06258 (2007).

21 Burgess, S., Dudbridge, F. & Thompson, S. G. Combining information on multiple instrumental variables in Mendelian randomization: comparison of allele score and summarized data methods. Stat. Med. 35, 1880–1906 (2015).

22 Burgess, S., Foley, C. N. & Zuber, V. Inferring Causal Relationships Between Risk Factors and Outcomes from Genome-Wide Association Study Data. Annu Rev Genomics Hum Genet 19, 303–327, doi:10.1146/annurev-genom-083117-021731 (2018).

23 Hemani, G., Bowden, J. & Davey Smith, G. Evaluating the potential role of pleiotropy in Mendelian randomization studies. Hum. Mol. Genet. 27, R195–R208, doi:10.1093/hmg/ddy163 (2018).

24 Lawlor, D. A., Harbord, R. M., Sterne, J. A., Timpson, N. & Davey Smith, G. Mendelian randomization: using genes as instruments for making causal inferences in epidemiology. Stat. Med. 27, 1133–1163, doi:10.1002/sim.3034 (2008).

25 Kamat, M. A. et al. PhenoScanner V2: an expanded tool for searching human genotype-phenotype associations. Bioinformatics 35, 4851–4853, doi:10.1093/bioinformatics/btz469 (2019).

26 Sanderson, E. Multivariable Mendelian Randomization and Mediation. Cold Spring Harb. Perspect. Med. 11, doi:10.1101/cshperspect.a038984 (2021).

27 Slob, E. A. W. & Burgess, S. A comparison of robust Mendelian randomization methods using summary data. Genet. Epidemiol. 44, 313–329, doi:10.1002/gepi.22295 (2020).

28 Ogasawara, H. Asymptotic standard errors of estimated standard errors in structural equation modelling. Br. J. Math. Stat. Psychol. 55, 213–229, doi:10.1348/000711002760554552 (2002).

29. R Core Team (2022). R: A language and environment for statistical computing. R Foundation for Statistical Computing, Vienna, Austria. URL https://www.R-project.org/.

30 Doughty, M. J. & Zaman, M. L. Human corneal thickness and its impact on intraocular pressure measures: a review and meta-analysis approach. Surv. Ophthalmol. 44, 367–408, doi:10.1016/s0039-6257(00)00110-7 (2000).

31 Holmberg, M. J. & Andersen, L. W. Collider Bias. JAMA 327, 1282–1283, doi:10.1001/jama.2022.1820 (2022).

32 Hernán, M. A. & Monge, S. Selection bias due to conditioning on a collider. BMJ 381, 1135, doi:10.1136/bmj.p1135 (2023).

