## Supplementary tables and figures for "Central corneal thickness and the risk of primary open-angle glaucoma: a Mendelian randomization mediation analysis"

Supplementary Figures and Tables

**Supplementary Figure 1** Funnel plot of single SNP Wald ratio estimates for the effect of central corneal thickness on primary open-angle glaucoma

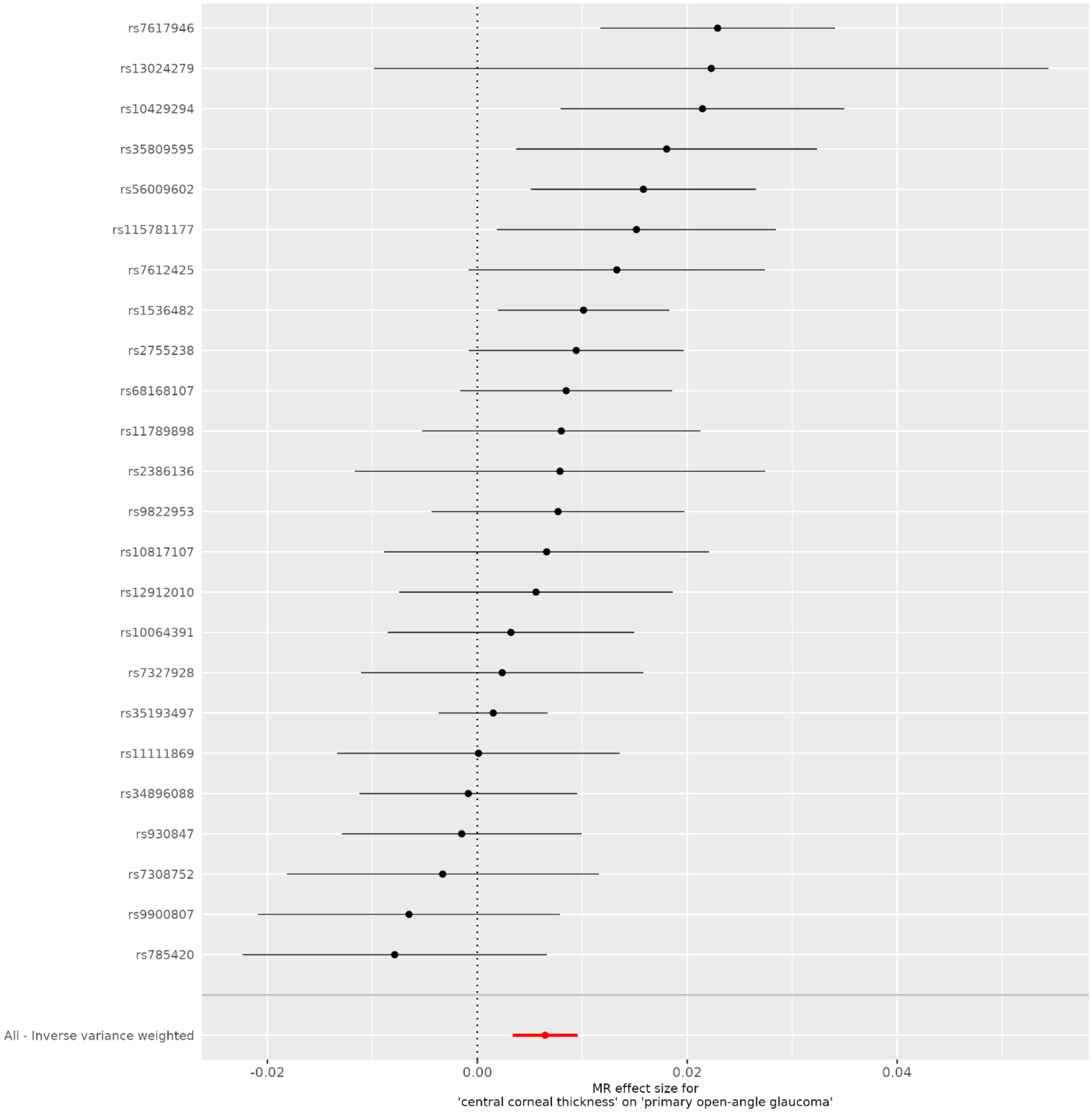


SNP: single-nucleotide polymorphism

**Supplementary Figure 2** Scatter plot of SNP-central corneal thickness associations vs SNP-primary open-angle glaucoma associations

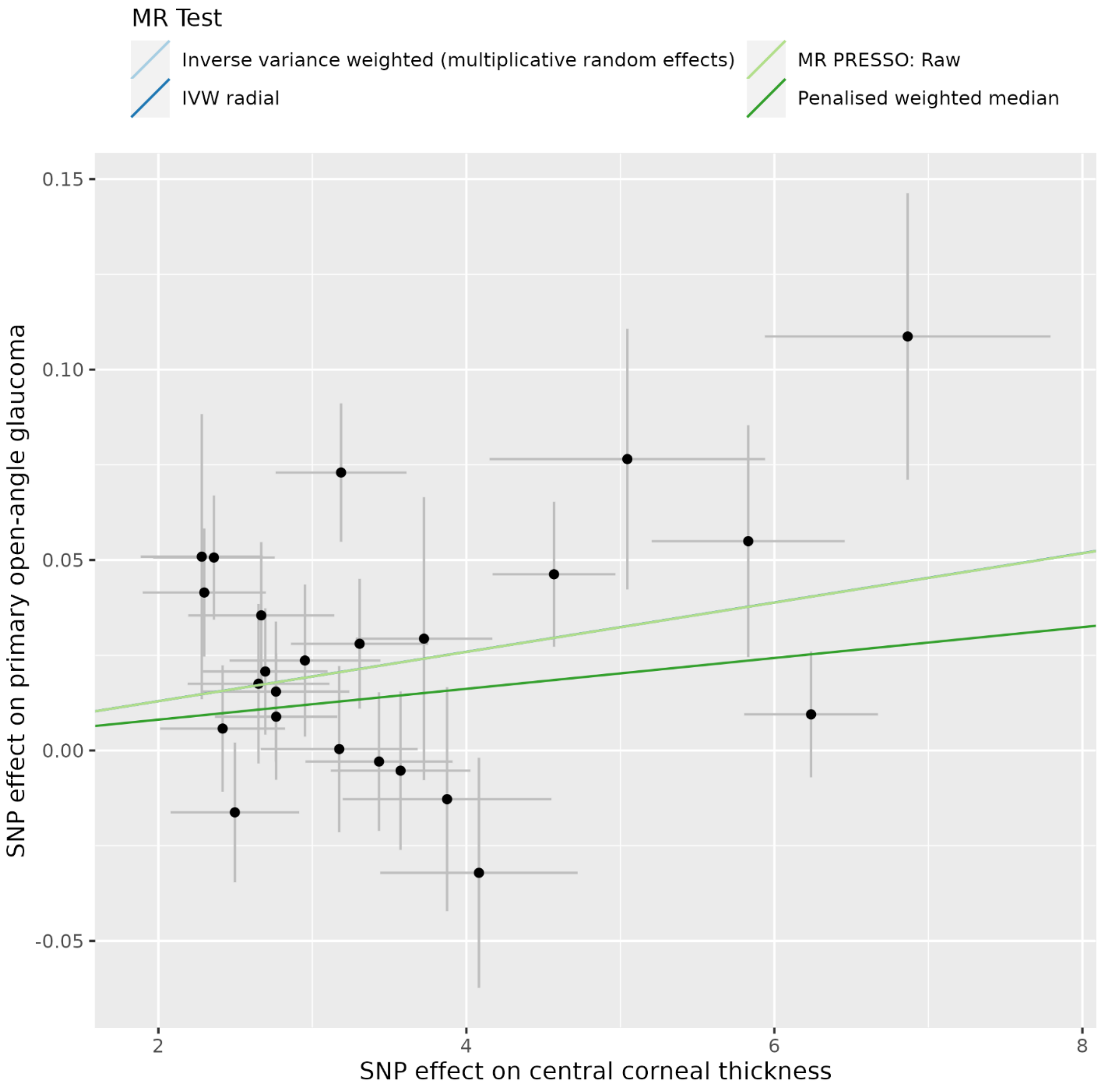


SNP: single-nucleotide polymorphism. Lines for inverse variance weighted and inverse variance weighted radial are not visible because they have the same slope with MR PRESSO line.

**Supplementary Table 1** Phenotypic descriptive statistics of studies included in the exposure, confounder/outcome risk factor, and outcome genome-wide association studies

| GWAS / Study | Phenotype | | | | | |
|---|---|---|---|---|---|---|
| Iglesias et. al. [1] | Central corneal thickness | | | | | |
| | N | Mean CCT | SD CCT | Range | Range age | Sex (male %) |
| BATS | 1153 | 544.3 | 35.00 | 381:679 | 5;90 | 56% |
| BMES | 1048 | 539.8 | 33.38 | 441.1; 662.2 | 49;86 | 57.73% |
| CROA TIA- Korcula | 848 | 558.0 | 36.72 | 468; 779.5 | 18;98 | 35.14% |
| CROA TIA- Split | 782 | 559.2 | 35.40 | 445.5; 663.5 | 18;85 | 39.45% |
| CROA TIA- Vis | 591 | 560.9 | 34.20 | 441; 661.5 | 25;86 | 40% |
| EPIC | 184 | 558.5 | 39.60 | 454; 662 | 52;88 | 47.83% |
| GHS1 | 2727 | 551.4 | 34.60 | 442; 676 | 35;74 | 51.4% |
| GHS2 | 1118 | 562 | 34 | 412; 675 | 35;74 | 49.7% |
| Orcades | 1096 | 538 | 32.74 | 431.5; 663.0 | 18.11;89.21 | 37.59% |
| RS1 | 873 | 544.5 | 34.47 | 431.7; 678.5 | 56.5;96.26 | 50.76% |
| RS2 | 1215 | 547.6 | 34.18 | 443.0; 669.0 | 65.87;98.59 | 46.5% |
| RS3 | 2391 | 550.8 | 33.70 | 443.5; 674.0 | 51.53;89.87 | 43.44% |
| TEST | 686 | 544.3 | 35.00 | 381:679 | 5;90 | 56% |
| Twins UK | 2080 | 546.47 | 33.5 | 430.3; 657.5 | 16.11;83.26 | 1.78% |
| Raine | 1011 | 537.9 0 | 32.26 | 417;647 | 18;22 | 48.8% |
| | Primary open-angle glaucoma | | | | | |
| | N cases | N controls | | | | |
| FINNGEN [2] | 6785 | 349292 | | | | |
| UKBB [3] | 1498 | 404535 | | | | |
| | Intraocular pressure | | | | | |
| | N | | | | | |
| UKBB [3] | 97653 | | | | | |

**Supplementary Table 2** Associations of single nucleotide polymorphisms with central corneal thickness

| | | | Estimates for exposure | | | | | Estimates for primary open angle glaucoma | | | |
|---|---|---|---|---|---|---|---|---|---|---|---|
| SNP | EA | OA | EAF | BETA | SE | P | F | BETA | SE | P | N |
| Lifetime central corneal thickness | | | | | | | | | | | |
| rs10064391 | A | G | 0.629 | -2.765 | 0.397 | 3.2e-12 | 48.6 | -0.009 | 0.017 | 0.592 | 2 |
| rs10429294 | T | C | 0.504 | 2.361 | 0.395 | 2.2e-09 | 35.8 | 0.051 | 0.016 | 0.002 | 2 |
| rs10817107 | A | C | 0.214 | 2.650 | 0.460 | 8.6e-09 | 33.1 | 0.018 | 0.021 | 0.403 | 2 |
| rs11111869 | A | G | 0.170 | 3.174 | 0.510 | 4.8e-10 | 38.8 | 0.000 | 0.022 | 0.987 | 2 |
| rs115781177 | A | G | 0.930 | -5.045 | 0.894 | 1.7e-08 | 31.8 | -0.077 | 0.034 | 0.025 | 2 |
| rs11789898 | T | G | 0.184 | 2.952 | 0.490 | 1.7e-09 | 36.3 | 0.024 | 0.020 | 0.237 | 2 |
| rs12912010 | T | G | 0.221 | 2.764 | 0.476 | 6.4e-09 | 33.7 | 0.015 | 0.018 | 0.401 | 2 |
| rs13024279 | A | G | 0.436 | -2.282 | 0.396 | 8.5e-09 | 33.2 | -0.051 | 0.037 | 0.174 | 1 |
| rs1536482 | A | G | 0.337 | -4.569 | 0.398 | 2.0e-30 | 131.4 | -0.046 | 0.019 | 0.015 | 2 |
| rs2386136 | A | G | 0.495 | 3.725 | 0.443 | 4.4e-17 | 70.6 | 0.029 | 0.037 | 0.429 | 1 |
| rs2755238 | T | C | 0.899 | 5.830 | 0.626 | 1.3e-20 | 86.6 | 0.055 | 0.030 | 0.071 | 2 |
| rs34896088 | A | C | 0.791 | 3.433 | 0.478 | 6.5e-13 | 51.7 | -0.003 | 0.018 | 0.873 | 2 |
| rs35193497 | T | G | 0.358 | -6.238 | 0.434 | 8.6e-47 | 206.3 | -0.009 | 0.017 | 0.566 | 2 |
| rs35809595 | A | G | 0.405 | -2.298 | 0.400 | 9.0e-09 | 33.1 | -0.041 | 0.017 | 0.014 | 2 |
| rs56009602 | T | C | 0.050 | 6.866 | 0.927 | 1.3e-13 | 54.8 | 0.109 | 0.038 | 0.004 | 2 |
| rs68168107 | A | G | 0.265 | -3.307 | 0.446 | 1.2e-13 | 55.0 | -0.028 | 0.017 | 0.100 | 2 |
| rs7308752 | A | G | 0.910 | 3.875 | 0.677 | 1.1e-08 | 32.7 | -0.013 | 0.029 | 0.664 | 2 |
| rs7327928 | T | C | 0.684 | -2.417 | 0.406 | 2.5e-09 | 35.5 | -0.006 | 0.017 | 0.729 | 2 |
| rs7612425 | A | G | 0.762 | 2.668 | 0.473 | 1.7e-08 | 31.8 | 0.035 | 0.019 | 0.065 | 2 |
| rs7617946 | T | C | 0.732 | 3.186 | 0.425 | 6.6e-14 | 56.2 | 0.073 | 0.018 | 0.000 | 2 |

| | | | | | | | | | | | |
|---|---|---|---|---|---|---|---|---|---|---|---|
| rs785420 | A | T | 0.893 | 4.082 | 0.641 | 1.9e-10 | 40.6 | -0.032 | 0.030 | 0.288 | 2 |
| rs930847 | T | G | 0.770 | -3.573 | 0.453 | 3.2e-15 | 62.1 | 0.005 | 0.021 | 0.799 | 2 |
| rs9822953 | T | C | 0.670 | 2.694 | 0.404 | 2.6e-11 | 44.5 | 0.021 | 0.017 | 0.211 | 2 |
| rs9900807 | T | C | 0.286 | -2.497 | 0.417 | 2.1e-09 | 35.9 | 0.016 | 0.018 | 0.376 | 2 |

EA, effect allele. OA, other allele. EAF, effect allele frequency. SE, standard error. SNPs rs13024279 and rs2386136 were present only in the UKBB GWAS.

**Supplementary Table 3** Association ($P<5x10^{-8}$) of the single nucleotide polymorphisms used as instruments with confounders or outcome risk factors in PhenoScanner (accessed on 2023/08/17 using the phenoscanner function of the R MendelianRandomization package)

| SNP | Phenotypes | PMID |
|---|---|---|
| Lifetime cannabis use | | |
| rs10817107 | Monocyte count | 27863252 |
| rs11789898 | Eosinophil count | 27863252 |
| rs11789898 | Eosinophil percentage of granulocytes | 27863252 |
| rs11789898 | Eosinophil percentage of white cells | 27863252 |
| rs11789898 | High light scatter percentage of red cells | 27863252 |
| rs11789898 | High light scatter reticulocyte count | 27863252 |
| rs11789898 | Mean corpuscular hemoglobin | 27863252 |
| rs11789898 | Mean corpuscular volume | 27863252 |
| rs11789898 | Mean platelet volume | 27863252 |
| rs11789898 | Neutrophil percentage of granulocytes | 27863252 |
| rs11789898 | Platelet count | 27863252 |
| rs11789898 | Platelet distribution width | 27863252 |
| rs11789898 | Red cell distribution width | 27863252 |
| rs11789898 | Reticulocyte count | 27863252 |
| rs11789898 | Reticulocyte fraction of red cells | 27863252 |
| rs11789898 | Sum eosinophil basophil counts | 27863252 |
| rs11789898 | Platelet count PLT | 22139419 |
| rs11789898 | Platelet counts | 22139419 |
| rs12912010 | Allergic disease | 29083406 |

| | | |
|---|---|---|
| rs1536482 | Central corneal thickness | 20719862 |
| rs2755238 | Cornea | 20485516 |
| rs7617946 | Height | 25282103 |

PMID, PubMed ID. Body mass index was considered as a relevant confounder or risk factor for primary open angle glaucoma.

**Supplementary Table 4** Heterogeneity of Wald ratios and MR-Egger test for directional pleiotropy

| Central corneal thickness | Heterogeneity | | | |
|---|---|---|---|---|
| | Q | Degrees of Freedom | P | $I_{GX}^2$ |
| Central corneal thickness | 40.62 | 23 | 0.013 | 0.42 |
| | MR-Egger test for directional pleiotropy | | | |
| | Intercept | Standard error | P | |
| Central corneal thickness | 1.356e-02 | 0.017 | 0.438 | |

**Supplementary Table 5** Inverse variance weighted estimates in leave-one-out analysis in primary analysis

| SNP excluded | SNP | OR | (95% CI) | P value |
|---|---|---|---|---|
| Central corneal thickness | rs10064391 | 1.51 | (1.29;1.76) | <0.01 |
| | rs10429294 | 1.50 | (1.28;1.75) | <0.01 |
| | rs10817107 | 1.51 | (1.3;1.77) | <0.01 |
| | rs11111869 | 1.53 | (1.32;1.78) | <0.01 |
| | rs115781177 | 1.48 | (1.27;1.71) | <0.01 |
| | rs11789898 | 1.51 | (1.29;1.77) | <0.01 |
| | rs12912010 | 1.50 | (1.28;1.75) | <0.01 |
| | rs13024279 | 1.51 | (1.29;1.76) | <0.01 |
| | rs1536482 | 1.49 | (1.27;1.75) | <0.01 |
| | rs2386136 | 1.51 | (1.29;1.76) | <0.01 |
| | rs2755238 | 1.52 | (1.3;1.78) | <0.01 |
| | rs34896088 | 1.53 | (1.31;1.79) | <0.01 |
| | rs35193497 | 1.54 | (1.3;1.83) | <0.01 |
| | rs35809595 | 1.48 | (1.28;1.71) | <0.01 |
| | rs56009602 | 1.47 | (1.27;1.72) | <0.01 |
| | rs68168107 | 1.51 | (1.29;1.77) | <0.01 |
| | rs7308752 | 1.52 | (1.3;1.77) | <0.01 |
| | rs7327928 | 1.51 | (1.3;1.77) | <0.01 |
| | rs7612425 | 1.50 | (1.29;1.76) | <0.01 |
| | rs7617946 | 1.43 | (1.27;1.62) | <0.01 |
| | rs785420 | 1.53 | (1.31;1.78) | <0.01 |
| | rs930847 | 1.51 | (1.29;1.77) | <0.01 |
| | rs9822953 | 1.53 | (1.31;1.78) | <0.01 |
| | rs9900807 | 1.52 | (1.31;1.78) | <0.01 |

**Supplementary Table 6** Mediation effect of central corneal thickness on primary open-angle glacuoma via intra-ocular pressure changes.

| Exposure | Mediator | Outcome | Total effect of CCT on POAG | Effect of exposure (CCT) on mediator (IOP) | Effect of mediator (IOP) on outcome (POAG) | Mediation effect of CCT on POAG via IOP | | Mediated proportion |
|---|---|---|---|---|---|---|---|---|
| | | | Effect size (95% CI) | Effect size (95% CI) | Effect size (95% CI) | Effect size (95% CI) | P value | (%) (95% CI) |
| Central corneal thickness | Intraocular pressure | Primary open-angle glaucoma | 0.404 (0.240 to 0.568) | 0.060 (0.000 to 0.121) | 1.911 (1.552 to 2.271) | 0.115 (-0.003 to 0.233) | 0.055 | 28.4 (0 to 60) |

CCT: central corneal thickness; POAG: primary open-angle glaucoma; IOP: intraocular pressure

**References**


1. Iglesias, A. I. *et al.* Cross-ancestry genome-wide association analysis of corneal thickness strengthens link between complex and Mendelian eye diseases. *Nat Commun* **9**, 1864, doi:10.1038/s41467-018-03646-6 (2018).
2. Kurki, M. I. *et al.* FinnGen provides genetic insights from a well-phenotyped isolated population. *Nature* **613**, 508-518, doi:10.1038/s41586-022-05473-8 (2023).
3. Bycroft, C. *et al.* The UK Biobank resource with deep phenotyping and genomic data. *Nature* **562**, 203-209, doi:10.1038/s41586-018-0579-z (2018).
